# Poor Oral Health Is Associated with Worse Brain Imaging Profiles

**DOI:** 10.1101/2023.03.18.23287435

**Authors:** Cyprien A. Rivier, Daniela Renedo, Adam de Havenon, Thomas M. Gill, Sam Payabvash, Kevin N. Sheth, Guido J. Falcone

## Abstract

**Importance:** Poor oral health is a modifiable risk factor that is associated with a variety of health outcomes. However, the relationship between oral and brain health is not well understood.

**Objective:** To test the hypothesis that poor oral health is associated with worse neuroimaging brain health profiles in persons without stroke or dementia.

**Design:** We conducted a 2-stage cross-sectional neuroimaging study using data from the UK Biobank (UKB). First, we tested for association between self-reported poor oral health and MRI neuroimaging markers of brain health. Second, we used Mendelian Randomization (MR) analyses to test for association between genetically-determined poor oral health and the same neuroimaging markers.

**Setting:** Ongoing population study in the United Kingdom. The UKB enrolled participants between 2006 and 2010. Data analysis was performed from September 1, 2022, to January 10, 2023.

**Participants:** 40,175 persons aged 40 to 70 enrolled between 2006 to 2010 who underwent a dedicated research brain MRI between 2012 and 2013.

**Exposures:** During MRI assessment, poor oral health was defined as the presence of dentures or loose teeth. As instruments for the MR analysis, we used 116 independent DNA sequence variants known to significantly increase the composite risk of decayed, missing, or filled teeth and dentures.

**Main Outcomes and Measures:** As neuroimaging markers of brain health, we assessed the volume of white matter hyperintensities (WMH), as well as aggregate measures of fractional anisotropy (FA) and mean diffusivity (MD), two metrics indicative of white matter tract disintegrity obtained through diffusion tensor imaging. These measurements were evaluated across 48 distinct brain regions, with FA and MD values for each region also considered as individual outcomes for the MR method.

**Results:** Among study participants, 5,470 (14%) had poor oral health. We found that poor oral health was associated with a 9% increase in WMH volume (beta = 0.09, standard deviation (SD) = 0.014, p P< 0.001), a 10% change in the aggregate FA score (beta = 0.10, SD = 0.013, P < 0.001), and a 5% change in the aggregate MD score (beta = 0.05, SD = 0.013, P < 0.001). Genetically-determined poor oral health was associated with a 30% increase in WMH volume (beta = 0.30, SD = 0.06, P < 0.001), a 43% change in aggregate FA score (beta = 0.42, SD = 0.06, P < 0.001), and an 10% change in aggregate MD score (beta = 0.10, SD = 0.03, P = 0.01).

**Conclusions and Relevance:** Among middle age Britons without stroke or dementia enrolled in a large population study, poor oral health was associated with worse neuroimaging brain health profiles. Genetic analyses confirmed these associations, supporting a potential causal association. Because the neuroimaging markers evaluated in the current study are established risk factors for stroke and dementia, our results suggest that oral health may be a promising target for interventions focused on improving brain health.

## INTRODUCTION

Poor oral health is a significant public health problem that has been linked to a variety of health outcomes. Epidemiological studies indicate that poor oral health, specifically periodontitis and tooth loss, increases the risk of cognitive decline and dementia^1–3^. Additionally, tooth loss and periodontitis have been identified as risk factors for stroke^4–7^. However, these studies were observational in nature and only examined clinical outcomes, rather than biomarkers of brain health that precede clinically evident manifestations by many years.

White matter disruption, as evaluated by white matter hyperintensities (WMHs) and white matter architecture disorganisation, has been linked to higher risks of cognitive decline, dementia, and stroke^8–11^ and could mediate the effects of oral health on brain health. Hence, we hypothesized that poor oral health is associated with white matter injury. To test this hypothesis, we investigated the relationship between poor oral health and white matter injury through both epidemiological and genetic approaches. The latter approach allowed us to evaluate a potential causal relationship between poor oral health and white matter damage^12,13^.

### METHODS

#### Study design

We conducted a 2-stage study that combined epidemiologic and genetic analyses. In the first stage, we conducted a cross-sectional observational analysis of participants in the UK Biobank (UKB) who had consented to the neuroimaging assessment. The UKB is a population-based cohort study that enrolled 502 480 community-dwelling volunteers across the United Kingdom between 2006 and 2010. Comprehensive information on the recruitment and enrollment of UKB participants has been previously detailed^14^. Participants with a history of stroke or dementia were excluded. The second stage involved a two-sample Mendelian randomization (MR) study design, where the genetic information for the exposure (poor oral health) and the outcome (neuroimaging profiles) are derived from different studies. This study received approval from the North West Centre for Research Ethics Committee.

#### Exposure of interest

During the neuroimaging assessment, participants in the UK Biobank completed a touchscreen survey to provide information on their medical history and overall health. The presence of clinically poor oral health was defined as a positive response to at least one of the following questions: “Do you have loose teeth?” or “Do you use dentures?”. These questions were specifically selected from the UKB dental health survey as they have been shown to possess the highest genetic correlation with periodontitis and tooth loss, respectively^15^.

#### Outcome

The acquisition protocol of the MRI neuroimaging study and the methods used to derive the outcomes of interest have been described in detail elsewhere^16–19^. We considered three neuroimaging markers of white matter injury: WMH volume, fractional anisotropy (FA) and mean diffusivity (MD). FA and MD are measures of white matter integrity obtained via diffusion tensor imaging that, along with WMH, are validated predictors of both stroke and dementia^8–11^. WMH volume was natural-log transformed and normalized (by subtracting the mean and dividing by the standard deviation). To generate aggregate measures of FA and MD for the entire brain, we computed the first principal component of values obtained from 48 different brain regions and normalized the derived values. In secondary analyses, we examined the FA and MD values of each of these 48 brain regions separately.

#### Genetic instruments

To select our genetic instruments, we utilized data from the Gene-Lifestyle Interactions in Dental Endpoints (GLIDE) consortium’s Genome Wide Association Study (GWAS)^15^. We selected 116 common (minor allele frequency >5%, Supplemental Table 1) single nucleotide polymorphisms (SNPs) associated with a composite trait of dentures and decayed, missing or filled tooth surfaces at genome-wide levels (*p* < 5e^-8^). To ensure independence of these SNPs, we filtered out variants with an r2 value greater than 0.1. Palindromic SNPs were excluded from all analyses. We subsequently estimated the effect of the genetic instruments on the neuroimaging markers of interest by conducting single-SNP association tests for each outcome in the UKB.

#### Statistical analysis

Continuous variables were reported as mean with standard deviation (SD) or median with interquartile range (IQR) as appropriate, while categorical variables were presented as count and percentage. To examine the association between poor oral health and neuroimaging outcomes of brain health, we used multivariable linear regression models that adjusted for potential confounders, including age, sex, and race/ethnicity (model 1) and age, sex, cardiovascular risk factors (hypertension, hypercholesterolemia, diabetes, smoking and body mass index) and comorbidities (history of myocardial infarction and atrial fibrillation, Model 2). Our primary MR analysis used the inverse variance weighted (IVW) method. In secondary analyses, we tested for horizontal pleiotropy (the possibility that the effect of the instrument on the outcome of interest is exerted through a pathway other than poor oral health) using the Mendelian Randomization Pleiotropy Residual Sum and Outlier (MR-PRESSO) global test. To account for this phenomenon, we implemented the weighted median method and calculated the MR-PRESSO outlier-corrected estimate. When considering each of the 48 brain regions as individual outcomes, we used the IVW method and applied Bonferroni correction to the obtained p-values by multiplying them by a factor of 96 (48 brain regions x 2 diffusion metrics). All analyses were performed in R version 4.1.3 using the packages MendelianRandomization and MR-PRESSO^20,21^.

## RESULTS

The baseline characteristics of the studied population are presented in Table 1 (mean age 55 (SD 7.5), 21,323 (53.0%) were female). Among the 40,175 participants, 5,470 (14%) had clinically poor oral health, which included 3,806 (12%) individuals with dentures and 684 (1.7%) with loose teeth.

**Table 1.** Baseline characteristics of the study population.

|  |  | Total population<br>N = 40,175 | Poor oral health<br>NO<br>N = 34,705 | Poor oral health<br>YES<br>N = 5,470 | P-value |
| --- | --- | --- | --- | --- | --- |
| Demographics |  |  |  |  |  |
| Mean age (SD) |  | 54.96 (7.5) | 54.30 (7.5) | 59.17 (6.6) | <0.001 |
| Female sex (%) |  | 21,323 (53.1%) | 18,769 (54.1%) | 2,554 (46.7%) | <0.001 |
| White ethnicity (%) |  | 38,892 (97.3%) | 33,601 (97.3%) | 5,291 (97.4%) | 0.01 |
| Vascular risk factors |  |  |  |  |  |
| Hypertension |  | 7,497 (18.7%) | 6,021 (17.3%) | 1,476 (27.0%) | <0.001 |
| Hypercholesterolemia |  | 3,419 (8.5%) | 2,709 (7.8%) | 710 (13.0%) | <0.001 |
| Type II diabetes |  | 1,040 (2.6%) | 810 (2.3%) | 230 (4.2%) | <0.001 |
| Smoking | Previous | 13,137 (32.8%) | 10,730 (31.0%) | 2,407 (44.1%) | <0.001 |
|  | Current | 2,467 (6.2%) | 1,917 (5.5%) | 550 (10.1%) |  |
| BMI | >=25 | 17,159 (42.8%) | 14,664 (42.3%) | 2,495 (45.7%) | <0.001 |
|  | >=30 | 7,012 (17.5%) | 5,766 (16.6%) | 1,246 (22.8%) |  |
| Cardiovascular Comorbidities |  |  |  |  |  |
| Atrial fibrillation |  | 835 (2.1%) | 668 (1.9%) | 167 (3.1%) | <0.001 |
| History of myocardial infarction |  | 753 (1.9%) | 564 (1.6%) | 189 (3.5%) | <0.001 |
| Oral health phenotypes |  |  |  |  |  |
| Dentures |  | 4,954 (12.3%) | 0 (0.0%) | 4,954 (90.6%) | - |
| Loose teeth |  | 717 (1.8%) | 0 (0.0%) | 717 (13.1%) | - |
| Dentures or loose teeth |  | 5,470 (13.6%) | 0 (0.0%) | 5,470 (100.0%) | - |

### First stage: Epidemiological analyses of oral and brain health

We found evidence of a relationship between poor oral health and worse neuroimaging profiles of brain health. After adjusting for age, sex and race/ethnicity (Model 1, Table 2), the presence of poor oral health was significantly associated with a 9% increase in WMH volume (beta = 0.09, SD = 0.014, P < 0.001), a 10% change in aggregate FA scores (beta = 0.10, SD = 0.013, P < 0.001), and a 5% change in aggregate MD scores (beta = 0.05, SD = 0.013, P < 0.001). Of note, these results remained significant when implementing different modeling strategies (Table 2), including unadjusted models (*WMH*: beta = 0.46, SD = 0.016, P < 0.001; *FA*: beta = 0.41, SD = 0.016, P < 0.001; *MD*: beta = 0.42, SD = 0.016, P < 0.001) and multivariable models that also adjusted for vascular risk factors (*WMH*: beta = 0.09, SD = 0.014, P < 0.001; *FA*: beta = 0.10, SD = 0.013, P < 0.001; *MD*: beta = 0.05, SD = 0.013, P < 0.001).

**Table 2.** Multivariate liner regression results for poor oral health.

| **Model** | **No Covariates** |  | **Demographics**<br>**Model 1** |  | **Demographics + CVRF\***<br>**Model 2** |  |
| --- | --- | --- | --- | --- | --- | --- |
| **Outcome** | **Beta (SE)** | **P** | **Beta (SE)** | **P** | **Beta (SE)** | **P** |
| **WMH volume** | 0.46 (0.016) | <0.001 | 0.12 (0.014) | <0.001 | 0.09 (0.014) | <0.001 |
| **Fractional Anisotropy** | 0.41 (0.016) | <0.001 | 0.13 (0.013) | <0.001 | 0.10 (0.013) | <0.001 |
| **Mean Diffusivity** | 0.42 (0.016) | <0.001 | 0.07 (0.013) | <0.001 | 0.05 (0.013) | <0.001 |

\*Cardiovascular Risk Factors: hypertension, hypercholesterolemia, diabetes, smoking, BMI, history of myocardial infarction and atrial fibrillation

### Second stage: Mendelian Randomization analyses of oral and brain health

Several MR approaches revealed a positive association between genetically determined poor oral health and worse brain health neuroimaging profiles (Table 3). In the primary analysis using the IVW MR method, genetically determined poor oral health was associated with a 30% increase in WMH volume (beta = 0.30, SD = 0.06, P < 0.001), a 43% change in aggregate FA score (beta = 0.43, SD = 0.06, P < 0.001), and a 10% change in aggregate MD score (beta = 0.10, SD = 0.03, P < 0.01). The weighted median method, a more conservative analytical approach, yielded similar results. The MR-PRESSO global test suggested the presence of pleiotropy (p<0.01 for the 3 outcomes), but the MR-PRESSO outlier-corrected estimate remained highly significant for all outcomes. After identifying and removing the SNPs causing the pleiotropy, genetically determined poor oral health was still associated with a 37% increase in WMH volume (beta = 0.37, SD = 0.06, P < 0.001), a 45% change in aggregate FA score (beta = 0.45, SD = 0.06, P < 0.001), and an 8% change in aggregate MD score (beta = 0.08, SD = 0.03, P = 0.01). The distortion test confirmed the absence of pleiotropy in outlier-corrected estimates (distortion p-values: 0.33 for WMH, 0.80 for FA, 0.39 for MD). Secondary analyses evaluating each of the 48 brain regions separately indicated that genetically determined poor oral health was associated with FA or MD changes in most of the tracts, even after adjusting for multiple testing (Figure 1, Supplemental figures 1, 2, and 3), with the retrolenticular part of the internal capsule emerging as the region with the most significantly altered microarchitecture (right retrolenticular part FA P= 5×10^−15^; MD P=3×10^−20^; left retrolenticular part FA P=4×10^−11^; MD P=9×10^−17^).

**Table 3.**
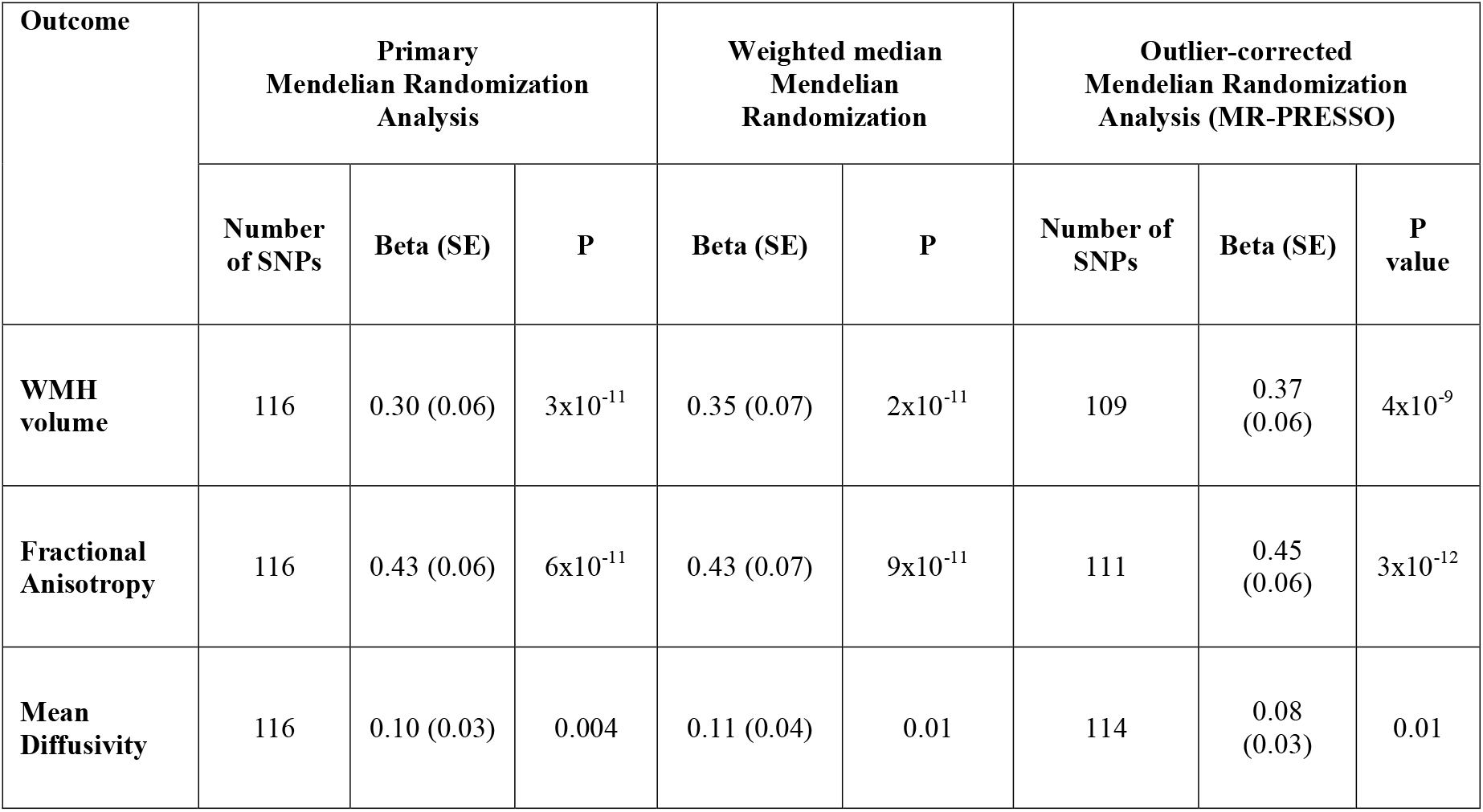
Mendelian Randomization analyses of genetically-determined oral health.

**Figure 1.**
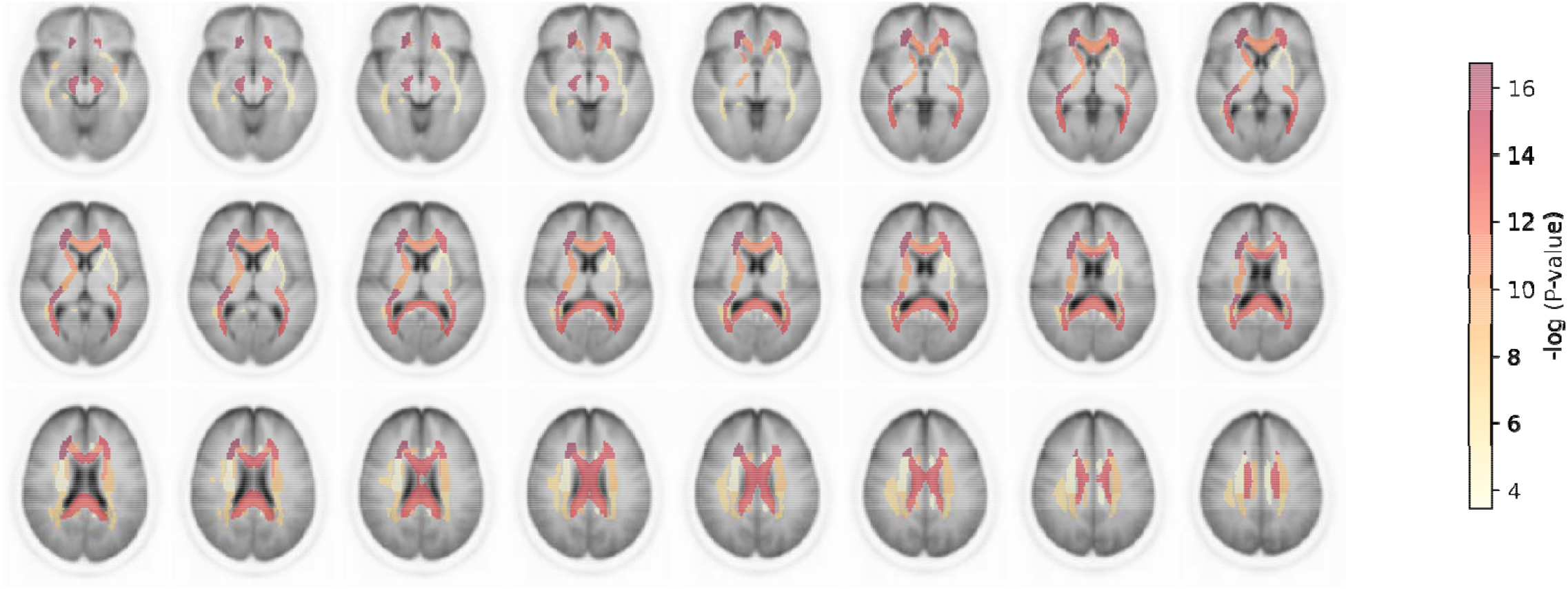
Statistical significance from Mendelian Randomization analyses between genetically-determined poor oral health and fractional anisotropy (FA) values across white matter (WM) tracts.

## DISCUSSION

The aim of our study was to investigate the association between poor oral health and neuroimaging markers of brain health. We employed a two-stage study design that included observational and genetic phases. In the observational analysis, we found that individuals with poor oral health had higher WMH volume and worse white matter architecture profiles, even after adjusting for confounding factors. Furthermore, our MR results confirmed these findings, thereby strengthening the case for potential causal associations^13,22^.

Previous epidemiologic studies found an association between poor oral health and higher risk of clinical outcomes related to brain health. Periodontitis and tooth loss have been linked to higher risk of cognitive decline^1,23^, while moderate or severe periodontitis has been identified as a risk factor for dementia^2^. Specifically, oral health in late adulthood has been found to be associated with the cognitive domains of learning and memory, complex attention, and executive control^24^. Periodontal treatment has also been shown to have a favorable effect on Alzheimer’s disease (AD)-related brain atrophy^25^, further supporting the link between oral and brain health. However, these studies were limited to cognitive outcomes that often become evident in older persons. Additionally, the purely observational nature of these studies precluded any causal inferences.

Our study adds important new evidence to this topic by showing that poor oral health leads to detrimental brain structure changes in persons without clinically-evident neurological disease. Of note, the neuroimaging markers of poor brain health used in our study are known to precede the onset of dementia and stroke, the two most important clinical entities related to brain health, by many years^8,10^. These findings support the notion that early intervention may carry important additional benefits. In addition, our study provides important new evidence indicating that the observed associations between oral and brain health are causal. Because genetic risk variants are randomly distributed during meiosis, variant-disease associations are relatively immune to confounding by post-partum confounders^12,26^. Along these lines, mounting evidence indicates that therapeutic interventions supported by genetic evidence in humans have twice the chances of being successful when evaluated in clinical trials^27^.

Further translational research is necessary to elucidate the neurobiological mechanisms linking poor oral health to brain health outcomes. One potential mechanism is systemic inflammation, which is commonly observed in patients with periodontal disease. Bacterial invasion of the gums can lead to the production of inflammatory mediators that enter the bloodstream, contributing to systemic inflammation^28^. Moreover, denture biofilm, containing a higher concentration of yeasts such as Candida albicans, has been implicated in inflammatory diseases like denture stomatitis^29^. Systemic inflammation, indicated by elevated CRP levels, has been associated with disruptions in white matter architecture^30,31^. Another possible mechanism involves tooth loss, a consequence of severe periodontal disease, leading to changes in dietary intake and diversity^32^. This could result in nutritional deficiencies, which are known risk factors for cognitive impairment and dementia^33^. Furthermore, the activity of the masticatory organ, linked to changes in the brain’s blood supply^34^, may also play a role. For instance, chewing exercises can help prevent Alzheimer’s disease by maintaining optimal blood flow to the brain^35^. Thus, tooth loss, by reducing masticatory organ activity, could decrease cerebrovascular blood flow and ultimately impact cognitive ability^23^. These findings hold significant implications for identifying modifiable risk factors for cognitive decline. Interventions focused on improving oral health may reduce the likelihood of developing cognitive impairment and dementia. However, this would require confirmation through clinical trials. Moreover, our results may inform public health policies aimed at promoting oral health and preventing adverse brain health outcomes at a population level.

In our exploratory MR analyses centered on tract-specific white matter architecture, we found that genetically determined poor oral health was linked to extensive microstructural damage in the majority of white matter tracts. These subtle alterations, indicative of demyelination or axonal degeneration, may manifest without visible macroscopic damage, acting as early indicators of underlying vascular pathology^36^. While we detected widespread damage in most tracts, the retrolenticular part of the internal capsule emerged as the region with the most robust association. This finding was observed in both the right and left sections of this brain area and was supported by both FA and MD values. DTI can identify microstructural abnormalities and axonal fiber damage before the development of WMH lesions^37^. Consequently, in future trials examining the impact of poor oral health treatment on brain health, the FA and MD values of this white matter tract could function as markers of treatment response prior to the emergence of WMH on structural MRI.

Our study has several strengths that reinforce the validity of our results. First, our observations were consistent across both the clinically and genetically determined exposures. Second, our findings revealed substantial effect size estimates, providing compelling evidence of a robust association between oral health and brain health. Additionally, we employed several conservative MR methods to assess the consistency of our results. Nevertheless, our study presents certain limitations that warrant consideration. Primarily, the UKB cohort is predominantly composed of individuals of European ancestry, potentially constraining the applicability of our findings to other populations. Furthermore, pleiotropy might affect our genetic analysis, although we employed various secondary analyses to mitigate this concern. Moreover, relying on self-reported measures to evaluate clinically poor oral health could introduce bias. Bearing these strengths and limitations in mind, our observations necessitate additional research and replication to conclusively establish a causal link between poor oral health and adverse brain health outcomes as determined by neuroimaging profiles.

In conclusion, our study offers important new evidence supporting an association between poor oral health and adverse neuroimaging brain health profiles. Using the Mendelian randomization technique, we provide strong support for a potential causal relationship between poor oral health and adverse neuroimaging markers of brain health. Confirmation of our findings through clinical trials is necessary to evaluate the potential impact of interventions to improve oral health on preventing cognitive impairment and dementia. Our findings have implications for public health policies, payer reimbursements, and social services. Additionally, they support the development of strategies that promote oral health as a viable means of maintaining brain health and preventing brain disease at the population level.

## Data Availability

All data produced in the present work are contained in the manuscript

## Acknowledgments

This research has been conducted using the UK Biobank Resource under Application Number 58743.

