## Supplemental Data for "Poor Oral Health Is Associated with Worse Brain Imaging Profiles"

**Supplemental Table 1. Single nucleotide polymorphisms included in the polygenic risk score.**

| CHR | BP | SNP | A1 | A2 | BETA | SE |
|---|---|---|---|---|---|---|
| 1 | 104028367 | rs79894207 | T | C | -0.017 | 0.003 |
| 1 | 104062750 | rs113922683 | A | G | -0.036 | 0.007 |
| 1 | 104364878 | rs72694438 | A | G | 0.021 | 0.003 |
| 1 | 155033572 | rs11264305 | A | G | -0.017 | 0.003 |
| 1 | 155155608 | rs4971099 | A | G | -0.021 | 0.003 |
| 1 | 210304319 | rs2046850 | T | C | -0.019 | 0.003 |
| 1 | 226868918 | rs3820640 | T | C | 0.02 | 0.004 |
| 2 | 417167 | rs62106258 | T | C | 0.042 | 0.006 |
| 2 | 630782 | rs13028737 | A | G | 0.019 | 0.003 |
| 2 | 25141538 | rs11676272 | A | G | -0.015 | 0.003 |
| 2 | 29616655 | rs80270335 | T | C | 0.027 | 0.005 |
| 2 | 69698158 | rs3552 | A | G | 0.016 | 0.003 |
| 2 | 155670203 | rs2652452 | A | C | -0.016 | 0.003 |
| 2 | 185921692 | rs263771 | A | C | 0.018 | 0.003 |
| 2 | 219327500 | rs10189064 | A | G | -0.047 | 0.007 |
| 2 | 219755011 | rs121908120 | A | T | -0.081 | 0.008 |
| 3 | 18852697 | rs9831002 | T | G | -0.016 | 0.003 |
| 3 | 25118637 | rs7429279 | A | C | 0.016 | 0.003 |
| 3 | 49971514 | rs7613875 | A | C | 0.018 | 0.003 |
| 3 | 136132705 | rs7620314 | A | G | -0.015 | 0.003 |
| 3 | 136443008 | rs61790808 | A | G | -0.016 | 0.003 |
| 3 | 193306987 | rs111289033 | A | G | -0.045 | 0.008 |
| 5 | 26928047 | rs55769264 | A | G | 0.015 | 0.003 |
| 5 | 44299998 | rs9292903 | A | G | -0.016 | 0.003 |
| 5 | 44539453 | rs1482698 | C | G | 0.02 | 0.003 |
| 5 | 107083487 | rs1352724 | A | C | -0.018 | 0.003 |
| 5 | 134374108 | rs4976261 | C | G | -0.019 | 0.003 |
| 5 | 134414688 | rs56353224 | T | C | 0.016 | 0.003 |
| 5 | 134492530 | rs17660141 | T | C | 0.042 | 0.005 |
| 5 | 134509987 | rs1122171 | T | C | 0.045 | 0.003 |
| 6 | 25174645 | rs7452710 | T | G | -0.016 | 0.003 |
| 6 | 25195517 | rs78712602 | T | C | 0.035 | 0.006 |
| 6 | 25464108 | rs301400 | T | C | -0.045 | 0.005 |
| 6 | 25758024 | rs9358885 | T | C | 0.02 | 0.003 |
| 6 | 25772639 | rs1892252 | C | G | -0.035 | 0.004 |
| 6 | 26008487 | rs9356996 | A | C | -0.02 | 0.003 |

| | | | | | | |
|---|---|---|---|---|---|---|
| 6 | 26037601 | rs13210041 | A | G | 0.042 | 0.005 |
| 6 | 26332605 | rs9467711 | A | C | -0.041 | 0.004 |
| 6 | 26336696 | rs9366651 | T | G | -0.029 | 0.003 |
| 6 | 26360597 | rs9348711 | A | G | -0.018 | 0.003 |
| 6 | 26522675 | rs4343916 | A | T | -0.018 | 0.003 |
| 6 | 26739487 | rs35144506 | A | G | -0.04 | 0.004 |
| 6 | 27075483 | rs9348752 | T | C | 0.024 | 0.004 |
| 6 | 27091661 | rs66462181 | T | C | -0.042 | 0.004 |
| 6 | 27491299 | rs6904596 | A | G | 0.043 | 0.004 |
| 6 | 27835218 | rs17763089 | A | G | 0.043 | 0.004 |
| 6 | 28130450 | rs34765154 | A | G | 0.043 | 0.004 |
| 6 | 28394680 | rs13201681 | T | C | 0.044 | 0.004 |
| 6 | 28665362 | rs7382146 | T | C | 0.016 | 0.003 |
| 6 | 28671343 | rs6908726 | C | G | -0.043 | 0.004 |
| 6 | 28983274 | rs3131082 | C | G | 0.043 | 0.004 |
| 6 | 29346329 | rs9257805 | A | G | -0.041 | 0.004 |
| 6 | 29607101 | rs3131856 | T | C | -0.038 | 0.004 |
| 6 | 29745439 | rs2394685 | A | G | 0.015 | 0.003 |
| 6 | 29905437 | rs3098023 | A | T | -0.038 | 0.004 |
| 6 | 30081108 | rs9261425 | T | C | 0.019 | 0.003 |
| 6 | 30138162 | rs1029239 | C | G | -0.015 | 0.003 |
| 6 | 30363136 | rs3094035 | T | G | 0.037 | 0.004 |
| 6 | 30607500 | rs9262131 | T | C | 0.015 | 0.003 |
| 6 | 30758857 | rs3129981 | T | C | 0.035 | 0.004 |
| 6 | 31081205 | rs2233956 | T | C | -0.03 | 0.003 |
| 6 | 31081838 | rs3130975 | T | C | 0.016 | 0.003 |
| 6 | 31312237 | rs4637682 | A | G | -0.017 | 0.003 |
| 6 | 31434621 | rs3131618 | A | G | -0.036 | 0.004 |
| 6 | 31591918 | rs2260051 | A | T | -0.015 | 0.003 |
| 6 | 31835164 | rs693906 | C | G | 0.035 | 0.004 |
| 6 | 32152442 | rs1800625 | A | G | -0.032 | 0.003 |
| 6 | 32221552 | rs9267995 | A | G | -0.015 | 0.003 |
| 6 | 32247990 | rs9268112 | A | G | -0.019 | 0.003 |
| 6 | 32473448 | rs111900528 | T | C | -0.019 | 0.003 |
| 6 | 32523109 | rs66988465 | T | C | 0.023 | 0.003 |
| 6 | 32525769 | rs187265184 | C | G | -0.025 | 0.003 |
| 6 | 32560341 | rs79966773 | T | C | -0.023 | 0.003 |
| 6 | 32604152 | rs9272324 | A | G | -0.019 | 0.003 |
| 6 | 32621132 | rs9366803 | T | C | -0.024 | 0.003 |

| | | | | | | |
|---|---|---|---|---|---|---|
| 6 | 32623550 | rs28430392 | A | C | -0.023 | 0.003 |
| 6 | 32625409 | rs28631719 | A | G | 0.015 | 0.003 |
| 6 | 32652281 | rs2856694 | T | C | -0.034 | 0.003 |
| 6 | 32754091 | rs4546530 | T | C | 0.017 | 0.003 |
| 6 | 32806391 | rs4148870 | T | C | 0.02 | 0.003 |
| 6 | 32946322 | rs76088152 | A | G | 0.027 | 0.005 |
| 6 | 33903427 | rs75808855 | T | C | 0.031 | 0.006 |
| 8 | 9229689 | rs898797 | T | C | 0.015 | 0.003 |
| 8 | 10240202 | rs9329221 | T | G | 0.015 | 0.003 |
| 8 | 10599994 | rs6982210 | A | G | -0.015 | 0.003 |
| 9 | 22542285 | rs10811723 | A | G | -0.019 | 0.003 |
| 9 | 79346204 | rs7852129 | A | C | -0.025 | 0.004 |
| 9 | 128661600 | rs10987008 | A | T | 0.021 | 0.003 |
| 10 | 10020194 | rs7918807 | T | C | 0.015 | 0.003 |
| 11 | 72444583 | rs7123876 | T | C | -0.017 | 0.003 |
| 11 | 72943483 | rs149467613 | A | G | -0.034 | 0.006 |
| 12 | 10704350 | rs10772314 | A | T | -0.015 | 0.003 |
| 15 | 63639416 | rs72748935 | T | C | -0.028 | 0.003 |
| 15 | 63660529 | rs10851728 | A | G | -0.025 | 0.003 |
| 15 | 63874881 | rs7180729 | A | T | -0.022 | 0.004 |
| 15 | 73353175 | rs6495046 | C | G | -0.017 | 0.003 |
| 15 | 73608231 | rs12440576 | A | G | 0.015 | 0.003 |
| 15 | 78915864 | rs10851907 | A | G | 0.016 | 0.003 |
| 15 | 90014945 | rs2072693 | T | G | 0.014 | 0.003 |
| 16 | 29958216 | rs8054556 | A | G | 0.016 | 0.003 |
| 16 | 51211595 | rs1108343 | T | C | 0.016 | 0.003 |
| 16 | 86710660 | rs10048146 | A | G | -0.026 | 0.003 |
| 17 | 45669524 | rs3865314 | A | C | 0.015 | 0.003 |
| 17 | 45996788 | rs12949191 | A | T | -0.015 | 0.003 |
| 17 | 46635649 | rs9905793 | A | G | 0.027 | 0.005 |
| 17 | 68399112 | rs34559440 | T | C | -0.016 | 0.003 |
| 17 | 70338127 | rs7217268 | A | G | 0.016 | 0.003 |
| 17 | 79361332 | rs57067187 | T | C | 0.016 | 0.003 |
| 18 | 57924823 | rs28822480 | A | G | 0.021 | 0.003 |
| 19 | 18718846 | rs2238651 | T | C | 0.017 | 0.003 |
| 19 | 49220323 | rs11672900 | A | G | -0.02 | 0.003 |
| 20 | 7654373 | rs4816017 | A | G | -0.017 | 0.003 |
| 22 | 30240778 | rs9614075 | T | C | -0.021 | 0.004 |
| 22 | 30581998 | rs9614155 | C | G | -0.021 | 0.004 |

| | | | | | | |
|---|---|---|---|---|---|---|
| 22 | 45727565 | rs1569414 | T | G | -0.02 | 0.003 |
| 22 | 45830784 | rs6007032 | C | G | -0.018 | 0.003 |

**Supplemental Figure 1:** Statistical significance of Mendelian Randomization (MR) analyses between genetically-determined poor oral health and mean diffusivity (MD) values across white matter (WM) tracts.

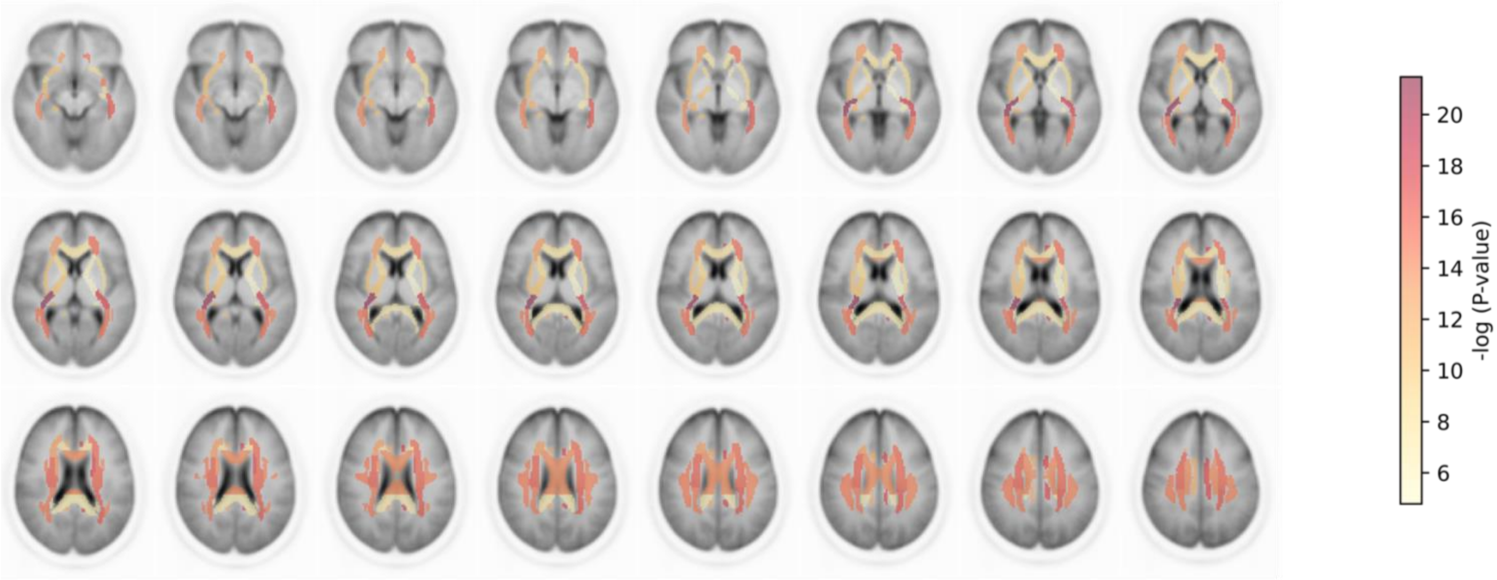

**Supplemental Figure 2:** Statistical significance and effect size of the MR analyses between genetically-determined poor oral health and fractional anistropy (FA) and mean diffusivity (MD) values across white matter (WM) tracts (part 1). P-values are Bonferonni-corrected for 96 simulatenous tests and values corresponding to p-values < 0.05 are not shown.

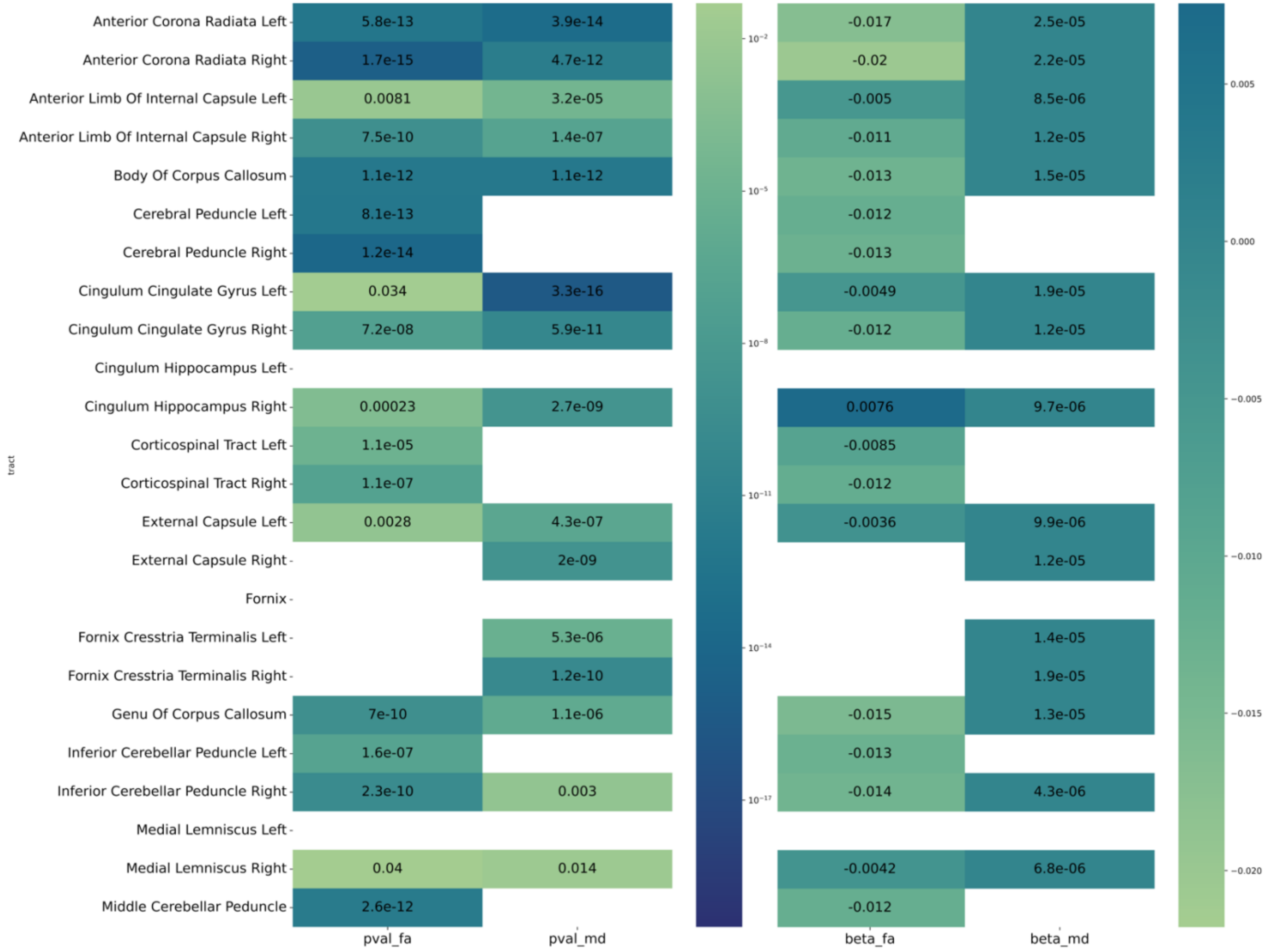

**Supplemental Figure 3:** Statistical significance and effect size of the MR analyses between genetically-determined poor oral health and fractional anistropy (FA) and mean diffusivity (MD) values across the second half of white matter (WM) tracts (part 2). P-values are Bonferonni-corrected for 96 simulatenous tests and values corresponding to p-values < 0.05 are not shown.

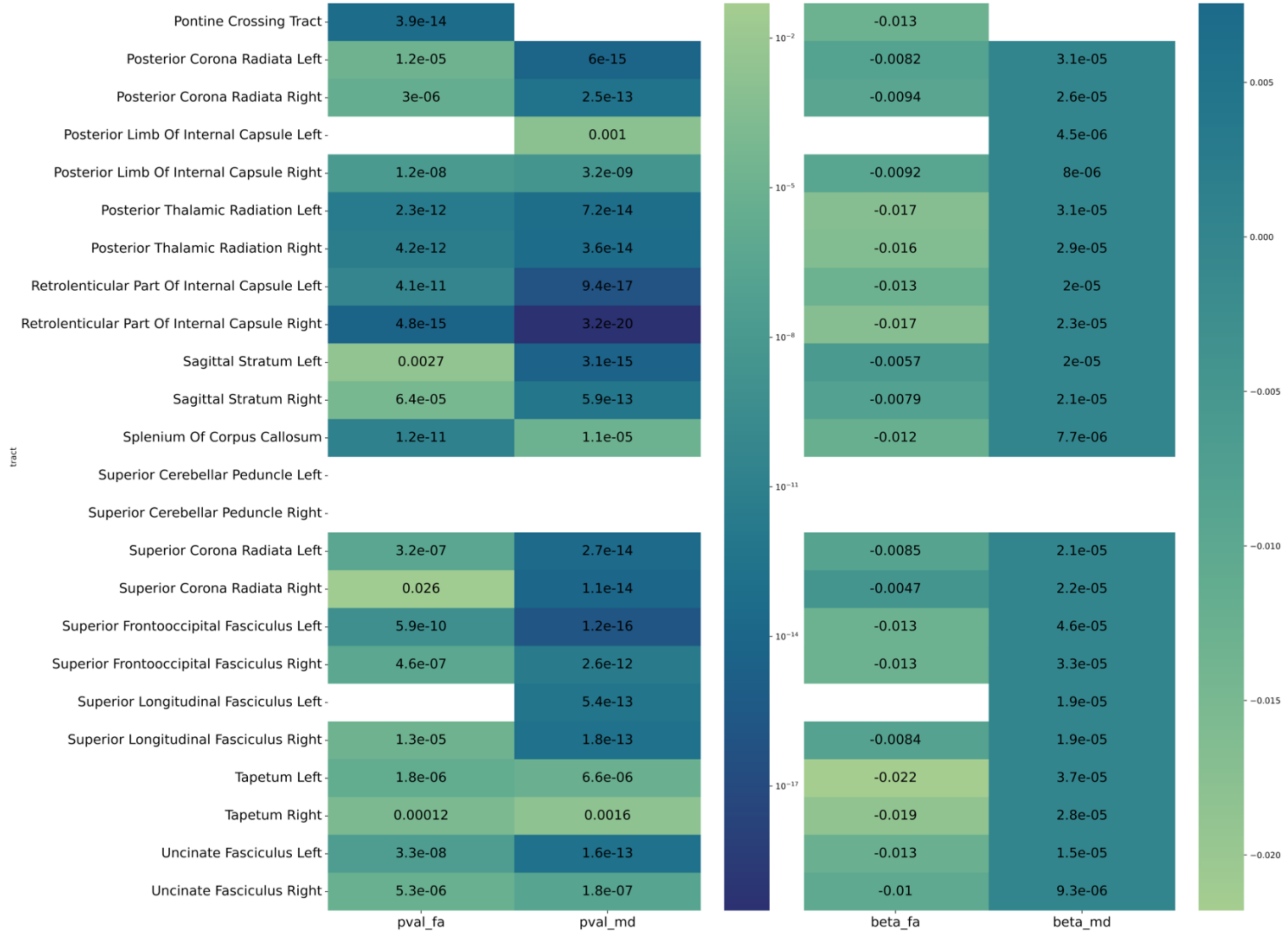
